# Evaluation of immunological responses to third COVID-19 vaccine among people treated with sphingosine receptor-1 modulators and anti-CD20 therapy

**DOI:** 10.1101/2022.06.10.22276253

**Authors:** Ilana Katz Sand, Sacha Gnjatic, Florian Krammer, Kevin Tuballes, Juan Manuel Carreño, Sammita Satyanarayan, Susan Filomena, Erin Staker, Johnstone Tcheou, Aaron Miller, Michelle Fabian, Neha Safi, Jamie Nichols, Jasmin Patel, Stephen Krieger, Stephanie Tankou, Sam Horng, Sylvia Klineova, Erin Beck, Miriam Merad, Fred Lublin

**Affiliations:** Corinne Goldsmith Dickinson Center for Multiple Sclerosis, Department of Neurology, Icahn School of Medicine at Mount Sinai, Icahn School of Medicine at Mount Sinai, New York, NY 10029; Precision Immunology Institute, Human Immune Monitoring Center, Icahn School of Medicine at Mount Sinai, New York, NY 10029; Department of Oncological Sciences, Icahn School of Medicine at Mount Sinai, New York, NY 10029; Department of Pathology, Icahn School of Medicine at Mount Sinai, New York, NY 10029; Department of Microbiology, Icahn School of Medicine at Mount Sinai, New York, NY, USA; Center for Vaccine Research and Pandemic Preparedness (C-VARPP), Icahn School of Medicine at Mount Sinai, New York, NY, USA

## Abstract

**Importance:** People living with multiple sclerosis (MS) and other disorders treated with immunomodulatory therapies remain concerned about suboptimal responses to coronavirus disease 2019 (COVID-19) vaccines. Important questions persist regarding immunological response to third vaccines, particularly with respect to newer virus variants.

**Objective:** Evaluate humoral and cellular immune responses to third COVID-19 vaccine dose in people on anti-CD20 therapy and sphingosine 1-phosphate receptor (S1PR) modulators, including Omicron-specific assays.

**Design:** Observational study evaluating immunological response to third COVID-19 vaccine dose in volunteers treated with anti-CD20 agents, S1PR modulators, and healthy controls. Neutralizing antibodies against USA-WA1/2020 (WA1) and B.1.1.529 (BA.1) severe acute respiratory syndrome coronavirus 2 (SARS-CoV-2) were measured before and after third vaccine. Cellular responses to spike peptide pools generated from WA1 and BA.1 were evaluated.

**Setting:** Mount Sinai Hospital

**Participants:** People treated with anti-CD20 therapy or S1PR modulators and healthy volunteers

**Exposure:** Treatment with anti-CD20 therapy, S1PR modulator, or neither

**Main outcomes and measures:** Serum neutralizing antibodies and *ex vivo* T cell responses against SARS-CoV-2 antigens.

**Results:** This cohort includes 25 participants on anti-CD20 therapy, 12 on S1PR modulators, and 14 healthy controls. Among those on anti-CD20 therapy, neutralizing antibodies to WA1 were significantly reduced compared to healthy controls (ID50% GM post-vaccination of 8.1 ± 2.8 in anti-CD20 therapy group vs 452.6 ± 8.442 healthy controls, P<0.0001) and neutralizing antibodies to BA.1 were below the threshold of detection nearly universally. However, cellular responses, including to Omicron-specific peptides, were not significantly different from controls. Among those on S1PR modulators, neutralizing antibodies to WA1 were detected in a minority, and only 3/12 had neutralizing antibodies just at the limit of detection to BA.1. Cellular responses to Spike antigen in those on S1PR modulators were reduced by a factor of 100 compared to controls (median 0.0008% vs. 0.08%, p<0.001) and were not significantly “boosted” by a third injection.

**Conclusions and Relevance:** Participants on immunomodulators had impaired antibody neutralization capacity, particularly to BA.1, even after a third vaccine. T cell responses were not affected by anti-CD20 therapies, but were nearly abrogated by S1PR modulators. These results have clinical implications warranting further study.

## Introduction

Many people receiving treatment with immunomodulatory therapies, including those living with multiple sclerosis (MS), remain apprehensive about COVID-19 risk.

Among common therapies for MS, there is high concern with anti-CD20 agents and sphingosine 1-phosphate receptor (S1PR) modulators. Anti-CD20 agents deplete circulating B cells and, to a lesser extent, subsets of CD4 and CD8 cells expressing low levels of CD20.^1^ In addition to widespread use in MS, anti-CD20 agents are utilized to treat B cell malignancies and a range of immune-mediated neurological and rheumatologic disorders.^2^ S1PR modulators functionally block S1PR function, sequestering lymphocytes in the lymph nodes and thymus.^3^ In addition to use in MS, the S1PR modulator ozanimod was recently approved for ulcerative colitis and ponesimod is currently in late stage clinical trials for psoriasis and graft-vs.-host disease.

Previous data suggest reduced humoral response to the primary vaccine series related to these treatments^4-6^. Limited evidence suggests preserved cellular response to vaccination with anti-CD20 therapies and reduced response with S1PR modulators.^7, 8^ Epidemiologic studies have suggested worse clinical outcomes with these therapies.^9^

Here we address the most recent significant questions regarding the production of neutralizing antibodies and cellular immune responses before and after third COVID-19 vaccinations in people treated with these immunomodulatory therapies, importantly accounting for the recent Omicron variant.

## Methods

### Sample

All participants were recruited between Mount Sinai’s MS Center and Human Immune Monitoring Center. Eligible participants began treatment with B cell or S1P modulator therapy at least 60 days prior to third vaccine or were healthy volunteers. For this analysis, we included all enrollees who had completed baseline sampling prior to a third injection and a follow-up sample approximately 4 weeks after the third vaccination by early January 2022.

### Assessing Humoral and Cellular Immunity

Microneutralization assays against USA-WA1/2020 (WA1, or wild-type) and B.1.1.529 (BA.1, or Omicron variant) SARS-CoV-2 were performed at the Icahn School of Medicine as previously described.^10^ Groups were compared by one-way ANOVA with Tukey multiple comparisons. T cell responses were assessed by IFN-gamma ELISPOT using 400,000 peripheral blood mononuclear cells pulsed with various antigens (15-mer overlapping peptide pool covering the entire sequence of Spike antigen (both WT and Omicron variants, from Miltenyi Biotec), N & Orf antigens (Bhardwaj, GenScript), long peptides covering the Spike RBD domain (Biosynthesis), recombinant Spike protein (gift from Drs. Herrera and Garforth, Einstein College of Medicine), and control viral peptide pools, DMSO, and PMA+ionomycine, as described.^11^ Responses were considered positive if >20 spots were detected and at least 2x more than the number of spots to DMSO. Pre-post comparisons were made by Wilcoxon paired t-tests, inter-cohort comparisons by Mann-Whitney t-test.

## Results

**Table 1** describes 25 participants on B cell therapy, 12 on S1PR modulators, and 14 controls.

**Table 1-.** Clinical Characteristics (n=51)

|  |  |  |  |  |
| --- | --- | --- | --- | --- |
| **Table 1 - Clinical Characteristics (n=51)** |  |  |  |  |
|  |  | Control (n=14) | S1P (n=12) | Bcell (n=25) |
| **Age (years), median (range)** |  | 31 (24-61) | 42 (31-69) | 49 (26-65) |
| **Sex** |  |  |  |  |
| Male, n (% total) |  | 9 (64%) | 7 (58%) | 12 (48%) |
| Female, n (% total) |  | 5 (36%) | 5 (42%) | 13 (52%) |
| **Race** |  |  |  |  |
| White, n (% total) |  | 12 (85%) | 11(92%) | 22 (88%) |
| Black, n (% total) |  | 0(0%) | 1 (8%) | 0 (0%) |
| Asian, n (% total) |  | 1 (7%) | 0 (0%) | 2 (8%) |
| Biracial, n (% total) |  | 1 (7%) | 0 (0%) | 1 (4%) |
| **Ethnicity** |  |  |  |  |
| Hispanic, n (% total) |  | 2 (14%) | 1 (8%) | 2 (8%) |
| Non-Hispanic, n (% total) |  | 12 (86%) | 11(92%) | 23 (92%) |
| **Disease Category** |  |  |  |  |
| none, n (% total) |  | 13(93%) | 0 (0%) | 0 (0%) |
| RRMS, n (% total) |  | 1 (7%) | 11 (92%) | 21 (84%) |
| SPMS, n (% total) |  | 0 (0%) | 1 (8%) | 1 (4%) |
| PPMS, n (% total) |  | 0 (0%) | 0 (0%) | 2 (8%) |
| MOG, n (% total) |  | 0 (0%) | 0 (0%) | 1 (4%) |
| **Disease Modifying Therapy (DMT)** |  |  |  |  |
| none, n (% total) |  | 13(93%) | 0 (0%) | 0 (0%) |
| teriflunomide, n (% total) |  | 1 (7%) | 0 (0%) | 0 (0%) |
| **S1P Class** |  |  |  |  |
| fingolimod, n (% total) |  | 0 (0%) | 8 (67%) | 0 (0%) |
| siponimod, n (% total) |  | 0 (0%) | 2 (17%) | 0 (0%) |
| ozanimod, n (% total) |  | 0 (0%) | 2 (17%) | 0 (0%) |
| **Anti-CD20 Class** |  |  |  |  |
| rituximab, n (% total) |  | 0 (0%) | 0 (0%) | 2 (8%) |
| ocrelizumab, n (% total) |  | 0 (0%) | 0 (0%) | 22 (88%) |
| ofatumumab, n (% total) |  | 0 (0%) | 0 (0%) | 1 (4%) |
| **DMT duration (days), median (range)** |  | n/a * | 1471 (69-3888) | 1185 (112-2629) |
| **Vaccine Type** |  |  |  |  |
| Pfizer, n (% total) |  | 11(79%) | 5 (42%) | 13 (52%) |
| Moderna, n (% total) |  | 3 (21%) | 5 (42%) | 12 (48%) |
| J&J, n (% total) |  | 0 (0%) | 2 (16%) | 0 (0%) |
| **Vaccine Interval\*\*** |  |  |  |  |
| days (median, range) |  | 279 (242-317) | 183 (147-257) | 207 (48-287) |
| **Immunological Testing** |  |  |  |  |
| T0 (pre-booster), days- (median, range) |  | 0 (0-7) | 5 (0-21) | 3 (0-21) |
| T4 (4 weeks post booster)- days (median, range) |  | 28 (26-45) | 28 (23-37) | 29 (22-39) |
| **Covid-19 Infection History\*\*\*** |  |  |  |  |
| None, n (% total) |  | 11 (79%) | 12 (100%) | 19 (76%) |
| Suspected, n (% total) |  | 2 (14%) | 0 (0%) | 3 (12%) |
| Confirmed, n (% total) |  | 1 (7%) | 0% | 3 (12%) |
| *DMT duration defined as time from DMT start to booster date for all patients. Of note, 1 control patient was on Aubagio (active comparator) and time from dmt start for that patient was 2694 days.<br>** Vaccine interval is defined by the time between the 2nd vaccination and 3rd vaccination for patients receiving Pfizer or Moderna vaccines, and is defined by time between 1st vaccination and 2nd vaccination for patients receiving J&J.<br>***Covid-19 Infection was defined as suspected or confirmed covid-19 prior to 3rd vaccination (or 2nd vaccination for patients with J&J). Controls (n=3), S1P (n=0), Anti-CD20 (n=6). |  |  |  |  |
|  | 8 |  |  |  |

**Figure 1** illustrates neutralizing antibody titers. Few on B-cell therapies mounted detectable neutralizing antibodies to WA1 (n=4, 2 just above the limit of detection (LoD)), with some increasing after the third vaccination (n=7, 4 just above the LoD). None had detectable neutralizing antibodies to BA.1 before the third vaccination, and only one converted after third vaccination. Among patients on S1PR modulators, three participants had neutralizing antibodies to BA.1 just at the level of detection post third injection. This contrasts with healthy controls, where a third vaccination resulted in detectable neutralizing antibodies to WA1 (ID50% GM of 59.25 ± 6.536 pre-vs 452.6 ± 8.442 post-vaccination, P<0.01) and BA.1 (ID50% GM of 6.13 ± 1.418 pre-vs 75.71 ± 3.405 post-vaccination, P<0.0001) nearly universally.

**Figure 1.**
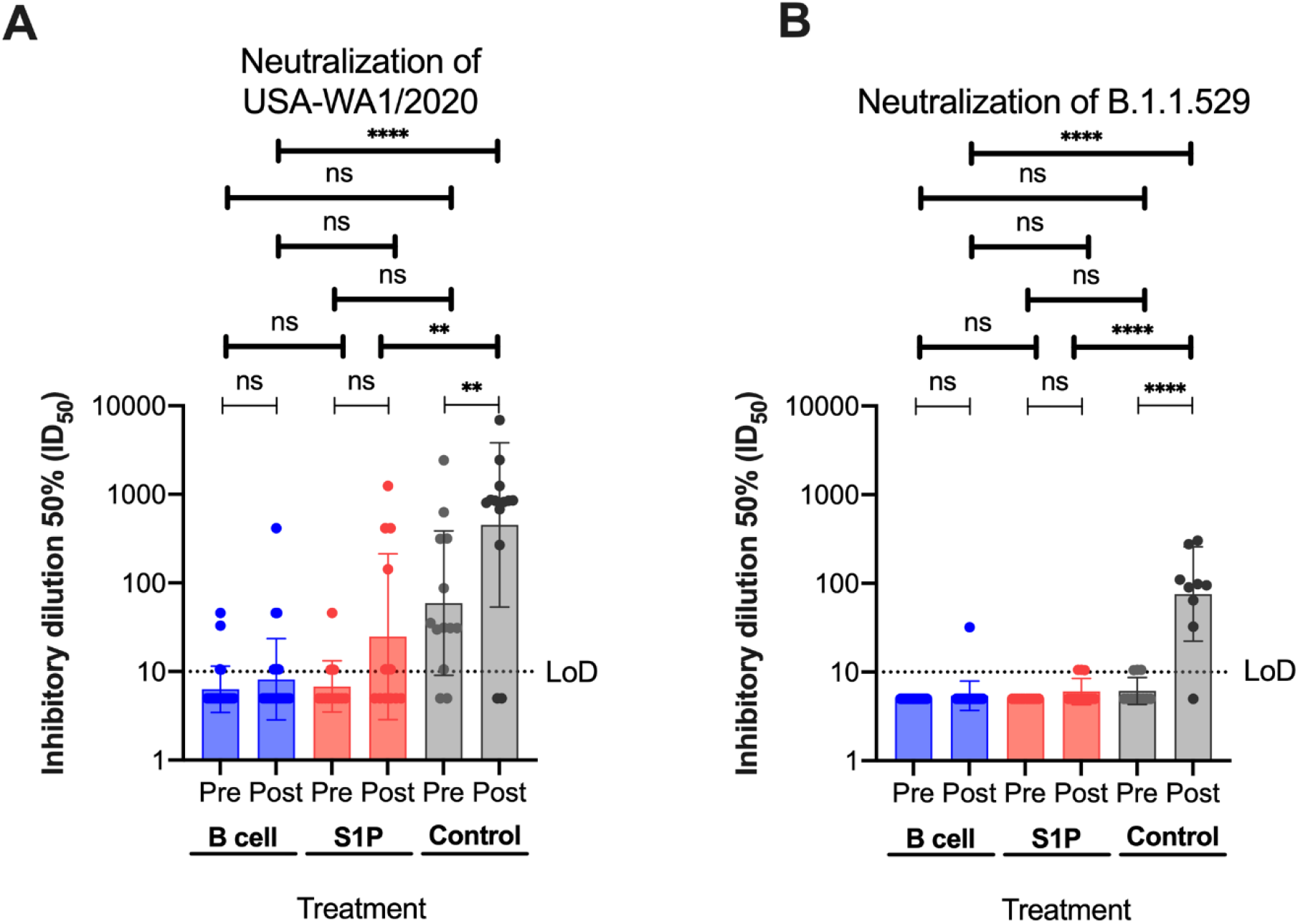
Neutralizing antibodies. Neutralization expressed as the inhibitory dilution 50% (ID_50_) of sera against USA-WA1/2020 (WT, shown in **A**) and B.1.1.529 (Omicron, shown in **B**) SARS-CoV-2 live viruses in the three groups: B cell, S1P, and healthy controls. Group comparisons performed using a One-way ANOVA with Tukey multiple comparisons; ns: not significant, * p<0.05, ** p<0.01, *** p<0.001, **** p<0.0001

**Figure 2** demonstrates the cellular response to spike peptide pools generated from WA1 **(A)** and BA.1 **(B)** SARS-CoV-2. Although there was more variability among those on B cell therapies compared to controls, overall T cell responses were not significantly different between B cell participants and controls. B cell participants showed significantly increased responses to WA1 and BA.1 peptides after a third injection (median 0.04% increase, p=0.008 by Wilcoxon paired test). Responses to WT Spike peptide pool were significantly greater in controls and B cell participants compared to S1PR participants (median 300-450 spots vs. 3 spots out of 400,000 PBMC, p<0.0001). Among S1PR participants, responses were not significantly different before and after third injection (median 0.004% increase). These results were confirmed with the Omicron peptide pool, RBD peptide pool, and Spike full-length protein, though these were generally less reactive in controls as well. Responses to the peptide pool generated from nucleocapsid (N) and open reading frame (ORF) proteins, which reflect naturally occurring immunity rather than vaccine-induced immunity, were not significantly different in MS participants compared to controls. Interestingly, responses to CEFT (cytomegalovirus, Epstein-Barr virus, influenza A virus, *Clostridium tetani*) peptide pools were significantly lower in B cell participants’ responses compared to controls.

**Figure 2.**
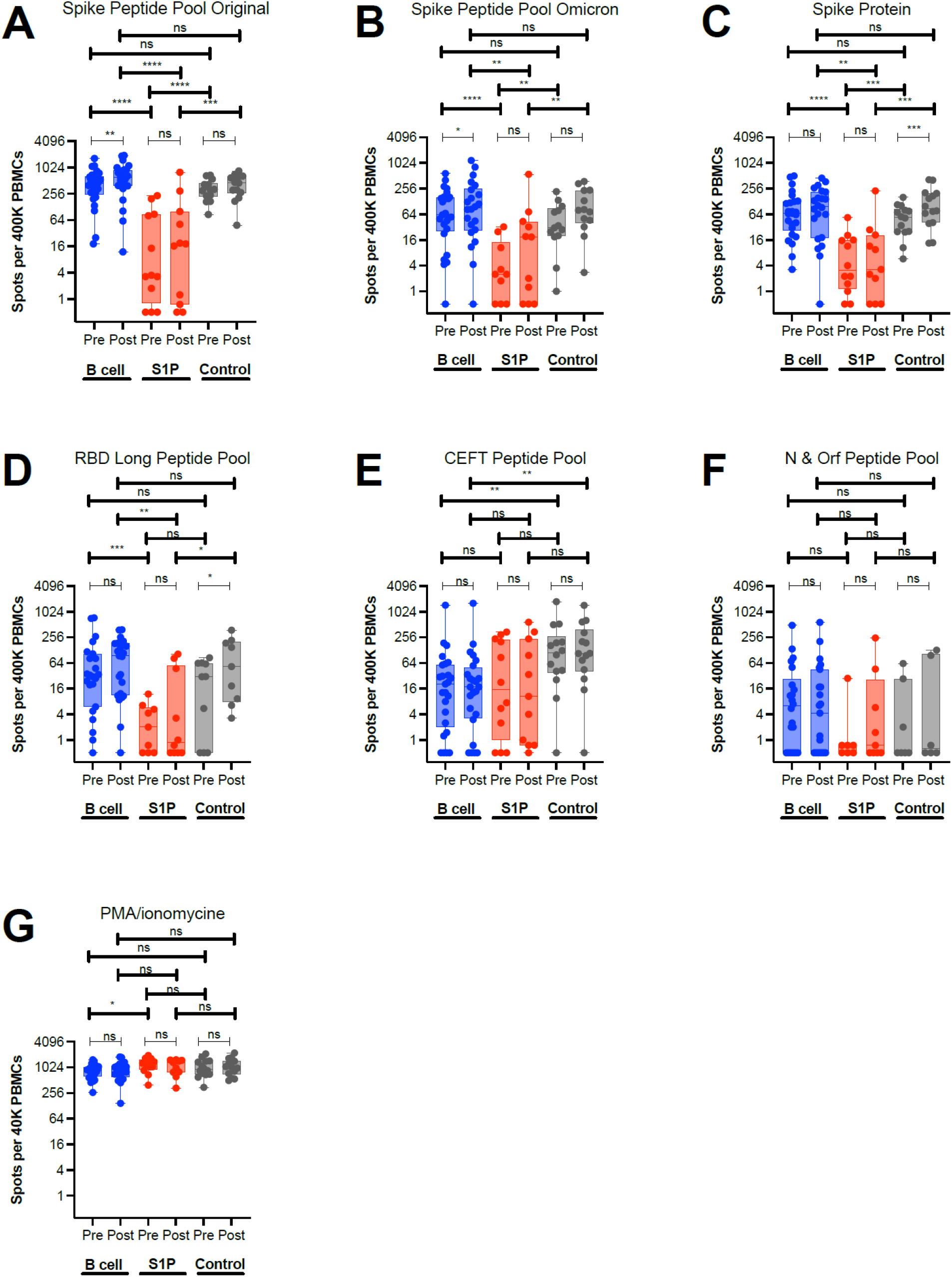
Cellular response. Number of positive interferon (IFN)-γ spots (log scale) out of 400,000 PBMC for the three groups: B cell, S1P, and healthy controls against the indicated antigens: Spike WT peptide pool WA1 (A); Spike omicron variant peptide pool (B); Spike full-length protein (C); long peptide pool covering Spike RBD domain (D); positive control viral peptide pool CEFT [from CMV, EBV, influenza, and tetanus] (E); SARS-CoV-2 N & Orf peptide pool (F); and assay control PMA/Ionomycin, out of 40,000 PBMCs (G). Intergroup comparisons by Mann-Whitney t-test in bold, pre-post comparisons by Wilcoxon paired test; ns: not significant, *: p<0.05, ** p<0.01, *** p<0.001, **** p<0.0001.

## Discussion

Participants on both therapies demonstrated significantly reduced production of neutralizing antibodies, particularly to BA.1, as compared to controls. Cellular immune responses to vaccination were similar among those on B cell therapies to controls and increased after a third vaccination. However, S1P participants demonstrated less robust cellular responses. Driving the lack of significant increase among S1P participants is that there were very few “converts” after the third vaccination. The cellular response increased with a third vaccination among those who had a measurable response to their first two injections, but did not increase for patients who never responded to the first two injections. This observation has significant consequences for expected responses to additional vaccines among those on S1PR modulators.

Timing of therapy initiation may play a role in vaccine response. The lone B cell participant with a high level of neutralizing antibodies to BA.1 after the third vaccination received the first vaccinations prior to starting therapy, though notably two others who started therapy after the first vaccinations did not develop neutralizing antibodies to BA.1 after the third vaccine. Of three S1P participants with neutralizing antibodies to BA.1 just at LOD, two received their first injections prior to starting therapy. These are important preliminary observations that will be relevant to those with new diagnoses or needing to change therapy going forward.

Strengths of our study include the presence of a control group and the study of S1PR modulators in addition to B cell therapy. We have assessed the levels of neutralizing antibodies, which reflect the effectiveness of the humoral immune response and represent a mechanistic correlate of protection against SARS-CoV-2,^12, 13^ and measured cellular immunity. We have also studied Omicron-specific assays, which more accurately reflect the current circulating strains.

Our study is limited by its small size. The cohort includes only one participant over age 65 because of the timing of enrollment (many of those over 65 had already received a third vaccine when enrollment began). We did not quantify the relative contribution of CD8 vs. CD4 T cell responses, though target antigen formulations used in our assays suggest that the third dose is better at increasing CD8 than CD4 T cells.^14^ Study design did not permit correlation to clinical outcomes.

Future directions include assessing durability of vaccine responses over time, studying the impact of natural infection with new variants, and most importantly, correlation with clinical outcomes. For those on S1PR modulators, future studies could consider brief cessation of therapy to aim for an improved response to additional injection, weighing potential benefits and risks of such a strategy.

## Data Availability

All data produced in the present study are available upon reasonable request to the authors and as allowed by institutional regulations

## Acknowledgments

The authors would like to thank the Fishman family for generously supporting this work, as well our patients at the CGD MS Center who graciously donated their time and samples to participate in this study.

SG was partially supported by NIH grants and contracts CA224319, DK124165, 75N91020R00055, CA263705, and CA196521.

Work in the Krammer laboratory was partially funded by the NIAID Collaborative Influenza Vaccine Innovation Centers (CIVIC) contract 75N93019C00051, the Centers of Excellence for Influenza Research and Surveillance (CEIRS, contract # HHSN272201400008C) and the Serological Sciences Network (SeroNet) in part with Federal funds from the National Cancer Institute, National Institutes of Health, under Contract No. 75N91019D00024, Task Order No. 75N91020F00003. The content of this publication does not necessarily reflect the views or policies of the Department of Health and Human Services, nor does mention of trade names, commercial products or organizations imply endorsement by the U.S. Government.

## References

1. Bar-Or A, O’Brien SM, Sweeney ML, Fox EJ, Cohen JA. Clinical Perspectives on the Molecular and Pharmacological Attributes of Anti-CD20 Therapies for Multiple Sclerosis. CNS Drugs. Sep 2021;35(9):985–997. doi:10.1007/s40263-021-00843-8

2. Lee DSW, Rojas OL, Gommerman JL. B cell depletion therapies in autoimmune disease: advances and mechanistic insights. Nat Rev Drug Discov. Mar 2021;20(3):179–199. doi:10.1038/s41573-020-00092-2

3. McGinley MP, Cohen JA. Sphingosine 1-phosphate receptor modulators in multiple sclerosis and other conditions. Lancet. Sep 25 2021;398(10306):1184–1194. doi:10.1016/s0140-6736(21)00244-0

4. Satyanarayan S, Safi N, Sorets T, et al. Differential antibody response to COVID-19 vaccines across immunomodulatory therapies for multiple sclerosis. Multiple Sclerosis and Related Disorders. doi:10.1016/j.msard.2022.103737

5. Tallantyre EC, Vickaryous N, Anderson V, et al. COVID-19 Vaccine Response in People with Multiple Sclerosis. Ann Neurol. Jan 2022;91(1):89–100. doi:10.1002/ana.26251

6. Sormani MP, Inglese M, Schiavetti I, et al. Effect of SARS-CoV-2 mRNA vaccination in MS patients treated with disease modifying therapies. EBioMedicine. Oct 2021;72:103581. doi:10.1016/j.ebiom.2021.103581

7. Gadani SP, Reyes-Mantilla M, Jank L, et al. Discordant humoral and T cell immune responses to SARS-CoV-2 vaccination in people with multiple sclerosis on anti-CD20 therapy. EBioMedicine. Nov 2021;73:103636. doi:10.1016/j.ebiom.2021.103636

8. Apostolidis SA, Kakara M, Painter MM, et al. Cellular and humoral immune responses following SARS-CoV-2 mRNA vaccination in patients with multiple sclerosis on anti-CD20 therapy. Nat Med. Nov 2021;27(11):1990–2001. doi:10.1038/s41591-021-01507-2

9. Sormani MP, Schiavetti I, Carmisciano L, et al. COVID-19 Severity in Multiple Sclerosis: Putting Data Into Context. Neurol Neuroimmunol Neuroinflamm. Jan 2022;9(1)doi:10.1212/nxi.0000000000001105

10. Carreño JM, Alshammary H, Tcheou J, et al. Activity of convalescent and vaccine serum against SARS-CoV-2 Omicron. Nature. 2022/02/01 2022;602(7898):682–688. doi:10.1038/s41586-022-04399-5

11. Sabbatini P, Tsuji T, Ferran L, et al. Phase I trial of overlapping long peptides from a tumor self-antigen and poly-ICLC shows rapid induction of integrated immune response in ovarian cancer patients. Clin Cancer Res. Dec 1 2012;18(23):6497–508. doi:10.1158/1078-0432.Ccr-12-2189

12. Khoury DS, Cromer D, Reynaldi A, et al. Neutralizing antibody levels are highly predictive of immune protection from symptomatic SARS-CoV-2 infection. Nature Medicine. 2021/07/01 2021;27(7):1205–1211. doi:10.1038/s41591-021-01377-8

13. Gilbert PB, Montefiori DC, McDermott AB, et al. Immune correlates analysis of the mRNA-1273 COVID-19 vaccine efficacy clinical trial. Science. 2022;375(6576):43–50. doi:doi:10.1126/science.abm3425

14. Gnjatic S, Atanackovic D, Matsuo M, et al. Cross-presentation of HLA class I epitopes from exogenous NY-ESO-1 polypeptides by nonprofessional APCs. J Immunol. Feb 1 2003;170(3):1191–6. doi:10.4049/jimmunol.170.3.1191

